# Prehabilitation before cancer surgery in the UK National Health Service: what services exist, and how do they address health inequalities?

**DOI:** 10.1101/2025.11.03.25338492

**Authors:** Hilary Stewart, Xiubin Zhang, Yasemin Hirst, Laura Wareing, Lisa Ashmore, Christopher Gaffney, Charlotte Hadley, Ces Kulikowski, Jo Rycroft-Malone, Cliff Shelton, Andrew Smith, PARITY Study Collaborators

## Abstract

**Objectives:** To identify and map prehabilitation services for patients preparing for cancer surgery in the UK National Health Service (NHS) and understand how issues of accessibility and inequality are being addressed.

**Design:** A survey based on the Template for Intervention Description and Replication and criteria for high-quality, equitable prehabilitation developed as part of our broader study.

**Setting:** National Health Service organisations in the UK

**Participants:** Representatives of prehabilitation services, who were mainly healthcare professionals. To be eligible for inclusion, services had to be part of the funded usual care pathway for patients in the NHS Trust or Health Board and offered, referred to, or signposted by the NHS cancer care team

**Outcomes:** The primary outcome was the availability of a prehabilitation service to patients who were preparing for cancer surgery at a given NHS organisation. Where services existed, we collected data on service characteristics, the size and workload of the service, screening and individualised assessment for prehabilitation, interventions, addressing inequalities, and evaluation of the service.

**Results:** Of 130 NHS organisations which provided cancer surgery services, we received a response from 112 (86%). Of these, 73 (65%) stated that they had an eligible prehabilitation service, and 39 (35%) stated that they did not. We received detailed survey responses from 51/73 services (70%). These demonstrated variability across all survey domains. Most services involved physiotherapists (43; 84%) and dietitians (37; 73%), with a variety of other professions represented. Twenty-four services (47%) reported that they tailor initial assessments to account for patient characteristics, and most services reported that they take steps to address inequities of access, the most common being support for people who have barriers to accessing the internet (46; 90%). Only 19 services (37%) were in receipt of permanent funding.

**Conclusions:** Prehabilitation provision for people preparing for cancer surgery varies widely across the UK, and this creates inequities in services. Nevertheless, prehabilitation services appear to be aware of the risk of unequal access, and are taking steps to address this. There is an opportunity to address inequalities as services are commissioned, developed and implemented.

**Strengths and Limitations of this Study:**

- This is the most comprehensive survey to date of prehabilitation services available to patients preparing for cancer surgery in the UK.
- Local collaborators helped us to identify services (or gaps in services) in 112 of 130 eligible organisations.
- Nevertheless, the localised and small-scale nature of prehabilitation services and inconsistent service naming conventions means we are likely to have missed some services.
- We identified important variability in service design, funding, and how services address health inequalities.
- While variations are identified, the methodology of survey-based research means that explanations for this variation remain speculative.

## Introduction

In the setting of cancer surgery, prehabilitation can be described as ‘the practice of enhancing a patient’s functional capacity before surgery, with the aim of improving postoperative outcomes’ [1]. Typically, it focuses on physical activity, diet, and psychological support, alone (unimodal) or in combination (multimodal) [2]. In recent years, prehabilitation has become an accepted component of many cancer surgery pathways in the UK and internationally [3-5].

A patient’s entry into a prehabilitation programme generally begins with a referral to a prehabilitation service, followed by screening for eligibility and an initial assessment where the level of prehabilitation provision is agreed. This is often classified into universal, targeted, and specialist provision, where universal prehabilitation is made available to all patients, and targeted and specialist provision are offered to those who are higher risk or have specific needs or comorbidities [2]. Targeted and specialist prehabilitation tends to involve more interventions, and more intensive input from more highly qualified or experienced staff. Although there are some larger, regional services, many are small in scale. This localised approach creates conditions for variation between regions, services and diagnoses, and therefore, the potential for disparities [6].

The evidence base for prehabilitation is characterised by heterogeneity across populations, interventions and outcome measures. In practice, what is delivered, how, and to whom, varies from service to service [6]. Because participation in prehabilitation requires engagement, time and access to facilities or resources, it may not be acceptable or accessible to all. Participation in prehabilitation is lower amongst people from socioeconomically deprived communities and some minority ethnic groups [7-10]. Studies also find that treatment and survival outcomes are poorer among these groups compared to socioeconomically advantaged and majority groups [10, 11]. This means that prehabilitation has the potential to exacerbate inequalities, because many of those who stand to benefit the most are less able to participate. The reasons for this are unclear, but it may stem from unequal access to exercise facilities, broadband internet, transport, and employment leave, for example.

Numerous service models for prehabilitation exist. While some are implemented at a healthcare system level, many are delivered in a localised fashion to specific patient cohorts. Different services are available to different people in different parts of the UK, and some areas have no services available at all. Following the COVID-19 pandemic, many local services were scaled back or stopped [12]. This heterogeneity of service provision is a further source of inequitable access, and effectively creates a ‘postcode lottery’ for patients [6].

To move towards more equitable provision of prehabilitation in the UK, it is essential to understand the current landscape, including what is already being done to address health inequalities. The purpose of this study was to address a gap in the evidence by identifying and mapping prehabilitation services for patients preparing for cancer surgery in the UK National Health Service (NHS) and understand how issues of accessibility and inequality are being addressed. This work provided foundational knowledge for of a wider mixed-methods study on quality and inequality in prehabilitation before cancer surgery (Prehabilitation Before Cancer Surgery: Quality and Inequality, abbreviated as ‘PARITY’, NIHR134282).

## Methods

We designed a survey based on the Template for Intervention Description and Replication (TiDIER) [13]. This is a checklist which includes specific items on the setting, funding, leadership and scope of a service. In addition to drawing on the TiDIER checklist, we integrated the findings of the initial part of our project, a Delphi study about the aims, objectives and values of prehabilitation [14], and consulted with our patient and public panel, to identify priority areas relevant to patient and professional stakeholders. These included items which explored access to services (e.g., travel, out-of-pocket expenses, digital literacy assessments), and governance arrangements (e.g., training, evaluation).

The survey was organised into seven sections: service characteristics, the size and workload of the service, screening and individualised assessment for prehabilitation, interventions, addressing inequalities, evaluation of the service, and key examples (optional). It was pilot tested by a prehabilitation service lead. A copy of the survey can be found in Supporting Information (Appendix A).

In line with the Macmillan definition of cancer prehabilitation services [2], NHS organisations across the UK meeting pre-specified criteria (Box 1) were eligible to take part in the survey. Services at all stages of implementation (including pilot services and services in development) were encouraged to participate.

### Box 1: Inclusion criteria

- Those with interventions designed to enhance a patient’s functional, nutritional and/or psychological capacity;
- Interventions delivered following a cancer diagnosis and prior to cancer surgery;
- Part of the funded usual care pathway for patients in the Trust / health board and offered, referred to, or signposted by the NHS cancer care team;
- May be for all cancer types and operations or specific cancers or operations;
- May be offered universally (all patients), targeted (high risk patients) or specialist (for those with complex needs);
- May be delivered in hospital, community, or online settings, including by a commissioned non-NHS provider (including third sector, commercial and local authorities).

Ethical approval was granted by the NHS Health Research Authority (Bradford Leeds Research Ethics Committee, IRAS ID 318939), and logged with the Lancaster University Faculty of Health and Medicine Research Ethics Committee.

The survey was administered using the Qualtrics XM survey platform (Qualtrics, Seattle, WA, USA) and delivered to participants by sharing a link with respondents via email. Recruitment ran from 9^th^ January – 31^st^ July 2024, and surveys closed 31^st^ August 2024. Reminders were sent to respondents to complete the survey at approximately 4-weekly intervals, no more than three times. Participants provided informed consent to participate as part of the questionnaire response, and this was recorded electronically.

Our initial approach to NHS organisations was based on contacting research and development (or equivalent) departments and requesting that our team be introduced to the prehabilitation service should one exist. This only yielded one response and, on enquiring about the reasons for this, it became clear that the term ‘prehabilitation’ is not consistently recognised in the naming of services, with some services favouring shorthand or informal terminology (prominent examples in the literature include ‘Prehab4Cancer’ in Manchester [5] and ‘Wesfit’ in Wessex [15]). Furthermore, prehabilitation services are hosted in a variety of departments (e.g., peri-operative medicine, physiotherapy) and may draw on staffing from multiple staff teams across an institution. In combination, these factors made it challenging for research and development departments to identify their services, and we therefore amended our protocol to allow us to engage with clinical contacts who possess local expertise.

We subsequently collaborated with existing networks (e.g. Trainees in Perioperative Medicine, (TRIPOM), Cancer Alliances) who contacted Trusts to establish whether they had a prehabilitation service accessible to patients preparing for cancer surgery. In addition, we advertised the call for participation at national prehabilitation events and via relevant professional organisations using a quick response (QR) code link. In areas of the UK with limited recruitment of respondents, the research team contacted Integrated Care Boards and Cancer Alliances/Networks to request further information regarding the presence or absence of services and to seek contact details of service leads. After confirming the presence of an eligible service, links to the survey were then shared with a service representative, who was typically the prehabilitation service lead.

Returned surveys were included in the analysis providing that at least 50% of the survey was completed, including completion of inequalities and service characteristics sections. Data were analysed descriptively, primarily focusing on the availability and accessibility of the services and reported nominally as well as using proportions. Where free-text responses were provided, these were analysed, taking a content analysis approach [16, 17]. Where we report free-text responses, these are summarised based on manifest content (i.e., explicitly and directly stated information) rather than latent content, which involves a deeper interpretation of data in context [16, 17].

### Patient and Public Involvement

The questionnaire was reviewed by a patient and public panel at the development stage, then refined in collaboration with a patient co-investigator (AP), with input from the panel. Patient and public involvement helped us to ensure that the questions were relevant to health inequalities and patient experiences.

## Results

There are approximately 229 NHS Trusts and Health Boards (hereafter, ‘NHS organisations’) in the UK. At the time of developing the survey, we identified 130 organisations that were likely to provide acute services such as cancer surgery (i.e., by excluding organisations such as ambulance Trusts, mental health Trusts and community Trusts) [18]. We received a response from 112 of these: 73 confirmed the presence of a prehabilitation service provided to patients before cancer surgery, and of these, 51 services completed a survey (see Table 1 for participant characteristics). Thirty-nine organisations confirmed there was no service, although two of these highlighted that patients were sent to another trust for prehabilitation, and one stated there had previously been a pilot service but that the service had not been renewed due to lack of funding. One Trust mentioned that their service comprised a commercial online prehabilitation platform.

**Table 1:**
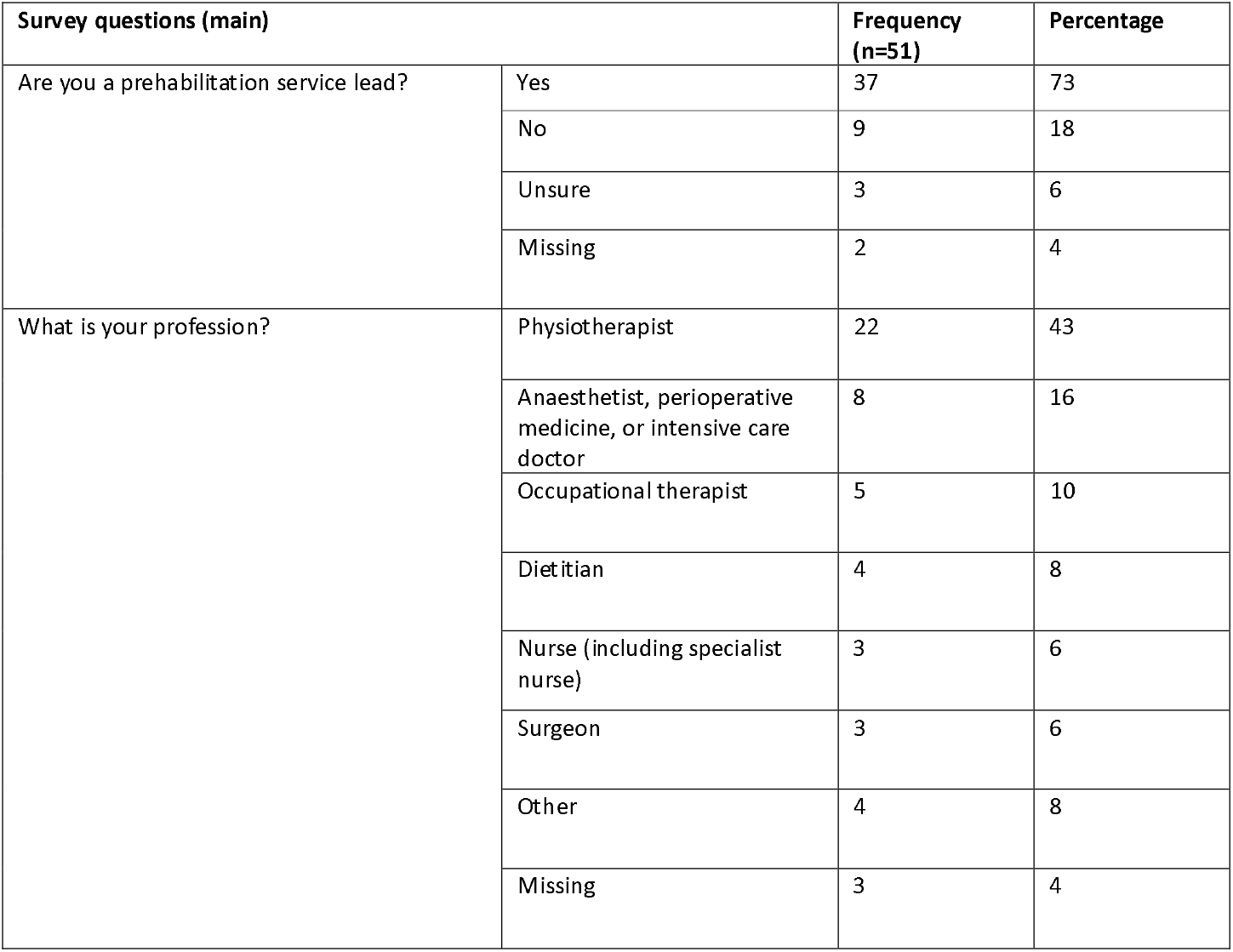
Characteristics of participants.

| Survey questions (main) |  | Frequency (n=51) | Percentage |
| --- | --- | --- | --- |
| Are you a prehabilitation service lead? | Yes | 37 | 73 |
|  | No | 9 | 18 |
|  | Unsure | 3 | 6 |
|  | Missing | 2 | 4 |
| What is your profession? | Physiotherapist | 22 | 43 |
|  | Anaesthetist, perioperative medicine, or intensive care doctor | 8 | 16 |
|  | Occupational therapist | 5 | 10 |
|  | Dietitian | 4 | 8 |
|  | Nurse (including specialist nurse) | 3 | 6 |
|  | Surgeon | 3 | 6 |
|  | Other | 4 | 8 |
|  | Missing | 3 | 4 |

All 51 survey responses contained sufficient data to be included in the analysis. They were from 48 NHS areas out of a total of 54 across England, Wales and Northern Ireland (there are 42 Integrated Care Boards in England, seven Health Boards in Wales, and five Health and Social Care Trusts in Northern Ireland). Broken down by nation, we received 43 responses from England (from 31 Integrated Care Boards), five from Wales and three from Northern Ireland (Fig. 1).

**Figure 1:**
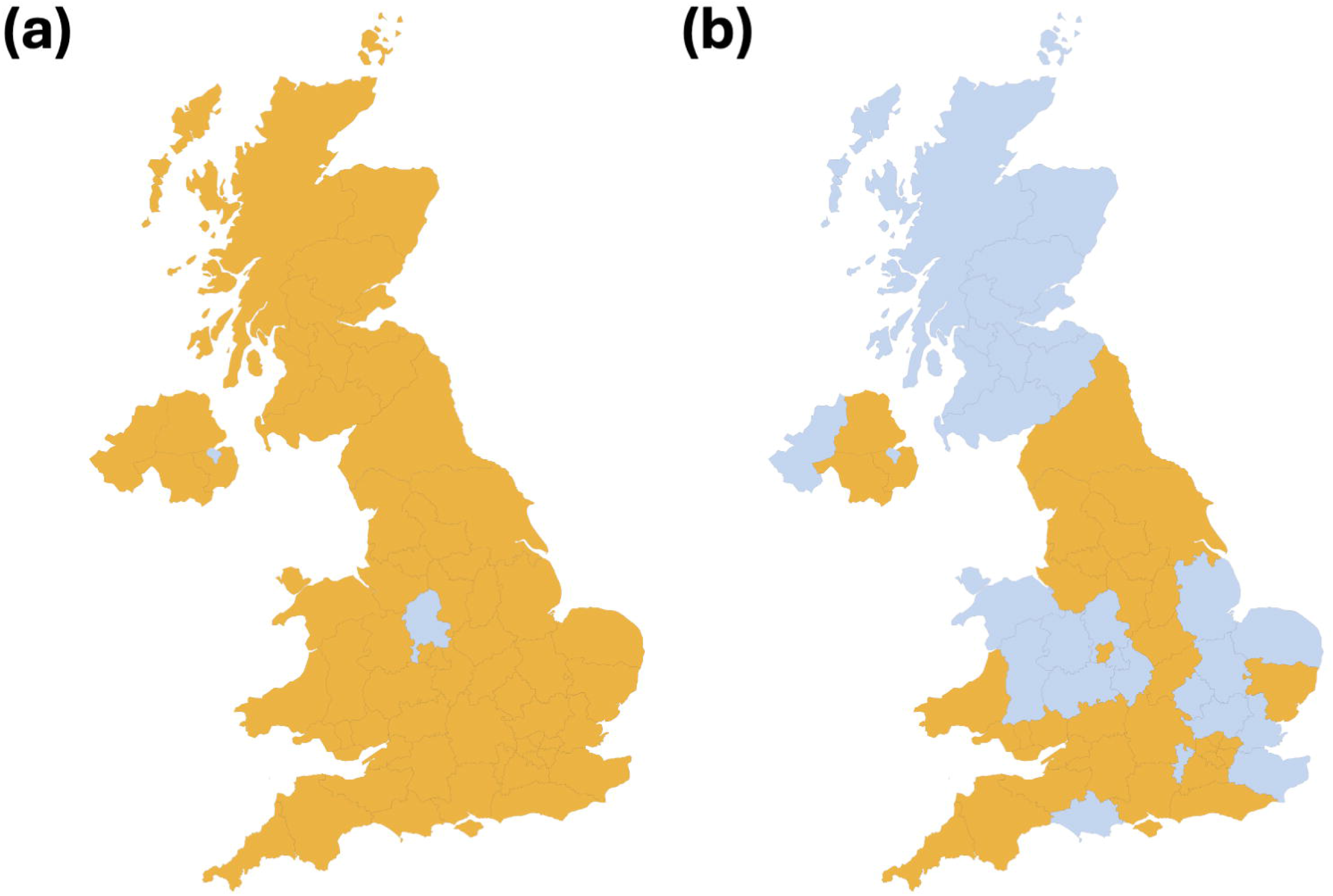
(a) geographical distribution (by NHS area) of responses to our survey; (b) geographical distribution of eligible services which returned a completed survey. Orange indicates a response, blue indicates no response.

Notably, no eligible services were identified in Scotland (which has 14 Health Boards) which fit our inclusion criteria. Following discussion with clinical contacts, we determined that this was due to the charity-based model adopted by most Scottish prehabilitation services at the time of the study, which maintained only informal links to NHS organisations, though contacts described that more formal prehabilitation service models were being developed.

Key findings on the characteristics of prehabilitation services are described below (more detailed information is available in Supporting Information (Appendix B).

Of the 51 positive responses to the survey, 46 (90%) provided a name for their prehabilitation service, 49 (96%) provided the name of their NHS organisation, while 48 (94%) named their hospitals. Of the services which provided their name, 28 services (55%) used ‘prehabilitation’ or ‘prehab’ in their name. Six services referred to activity and exercise in the context of cancer, and five services mentioned preparation for surgery. Ten services provided no name, and one of the respondents noted that due to not being substantively funded, they had no formal name.

Forty-one prehabilitation services (80%) were delivered by acute NHS organisations, confirming their role as the primary provider. Forty-eight (94%) of services indicated a known start date, with the earliest reported establishment of a prehabilitation service in 2014.

Forty-eight respondents (94%) specified the department within which their prehabilitation service was provided, and this indicated varied organisational structures; the most common department was physiotherapy (12 services; 24%).

Regarding coverage and accessibility, 26 services (51%) provide prehabilitation to patients from other hospitals within their organisation, while 14 (28%) do not. Nearly half (25; 49%) of respondents did not provide names of all the hospitals included in their prehabilitation service. Twenty-one services (41%) extended care to patients outside their area (e.g. Integrated Care Board), while 22 (43%) did not.

Nineteen services (37%) received permanent funding, and three services (6%) operated on a commissioned basis, while 23 (45%) operated as pilot programs with potential to extend, and five services (10%) remained unfunded. No services reported reliance on research grant funding.

Regarding staffing, the professions and specialties most involved in prehabilitation services were physiotherapists (43; 84%), dietitians (37; 73%), and cancer nurse specialists (22; 42%). Other professionals, such as anaesthetists (19; 37%), occupational therapists (17; 33%), and surgeons (14; 28%) also contributed. Eleven services (22%) involved a clinical psychologist, and three services (6%) involved a health psychologist.

It was uncommon for prehabilitation services to be available to all patients awaiting surgery for all cancer types. The most common cancer diagnoses which conferred eligibility for prehabilitation were lower gastrointestinal tract (37; 73%), lung (25; 49%), upper gastrointestinal tract (22; 43%), gynaecological (21; 41%), urological (20; 39%), hepatopancreatobiliary (16; 31%), head and neck (14; 28%), breast: (11; 22%) and haematological (10; 20%). Less common cancers included sarcomas (5; 10%), skin (5; 10%), and brain and central nervous system (3; 6%). No services reported providing care for children (age under 18) awaiting cancer surgery.

Regarding patient volume and eligibility, 36 services (71%) catered to fewer than 500 patients per year. Nine services (18%) catered for between 501-999 patients, and only two services (4%) supported more than 1000 patients per year. Thirty-two prehabilitation services (63%) extended to non-surgical oncology patients (e.g. undergoing chemotherapy or radiotherapy but not surgery), and 22 services (43%) also offered support to patients in receipt of palliative care. Fourteen services (28%) extended care to patient groups outside of cancer treatment (e.g., elective orthopaedic surgery).

Seventeen services (33%) reported that they had formal agreements with community-based or third-party providers, and 23 services (45%) maintained informal agreements. Thirty-four (67%) of respondents described that some or all of their service was provided by community based and third-party providers. The most common of these were exercise classes conducted at local leisure centres, led by a physiotherapist or a Personal Training (PT) instructor. Others included sessions organised by the local council, a university, or Macmillan Cancer Support (Supporting Information Appendix C includes more detailed data on referral and assessment processes).

Most respondents (43; 84%) reported having defined referral criteria for prehabilitation, while seven (14%) did not, and one (2%) did not answer. Screening and initial assessments were conducted at different appointments at 24 services (47%), while 21 (41%) carried them out at the same time. Five (10%) were unsure, and one (2%) did not respond.

Regarding referral sources, the primary referrers were cancer nurse specialists (46; 90%), followed by surgeons (27; 53%), anaesthetists (21; 41%), and oncologists (20; 39%). General practitioners were not reported to be involved in referrals (0%), while self-referrals were used in a small proportion of services (3; 6%), other sources, as indicated by respondents, accounted for referrals to 15 services (29%).

For those who referred patients to prehabilitation, face-to-face communication (42; 82%) was the most common method used when communicating with patients about what prehabilitation involves. Telephone communication (28; 55%) was also widely reported, while electronic communication (10; 20%) and mobile phone portals/apps (3; 6%) were less common. Letters (5; 10%) and other methods (8; 16%) are also used.

Most initial assessments took place in hospitals (31; 61%) and via telephone (27; 53%). Community centres (31; 26%) and home-based assessments (4; 8%) were less commonly utilised, while primary care was not used at all (0%). Other locations/methods were used by 12 services (24%).

Initial assessments were most frequently carried out by physiotherapists (41; 80%). Other professionals involved in initial assessment included dietitians (16; 31%), cancer specialist nurses (9; 18%), occupational therapists (9; 18%), and prehabilitation coordinators (6; 12%). While surgeons (1; 2%), anaesthetists (4; 8%), administration assistants (1; 2%), and nurses (1; 2%) played a comparatively minor role. Psychologists are not involved at all (0%). Twenty-two services (43%) indicated that “other” professionals are involved, as summarised in Box 2.

### Box 2: Other professionals involved in initial assessments

Cancer care coordinators

Exercise specialists (physiologists, technicians, therapists, instructors);

Prehabilitation support workers

Wellbeing practitioners

Clinical practitioners

Assistant/associate practitioners (physiotherapy, allied health professions and therapies)

Speech and language therapists

Nearly half of respondents (25; 49%) reported that assessments occurred at multiple points in patient trajectories, including shortly after diagnosis, after the decision for surgery, and before surgery. For others, assessments occurred shortly after cancer diagnosis (10; 20%), after the decision for surgery (11; 22%), or before surgery during pre-operative assessments (1; 2%). Three services (6%) reported no specified timing.

Regarding tailoring of initial assessments, responses were almost evenly split, with 24 (47%) stating that assessments are tailored to different patient groups (e.g., by sex, language), while 26 (51%) reported no tailoring. Free-text responses are summarised in Table 2.

**Table 2.**
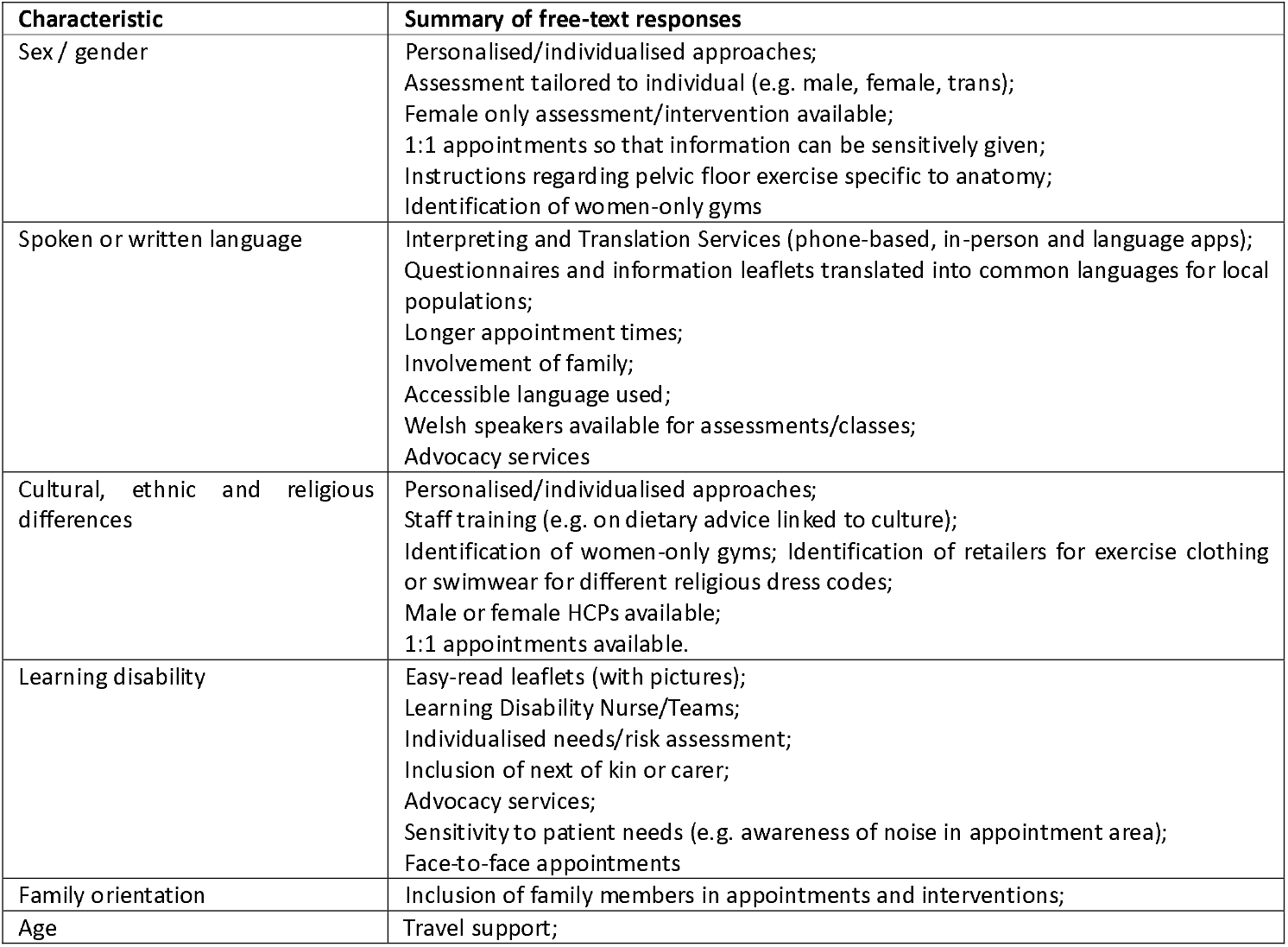
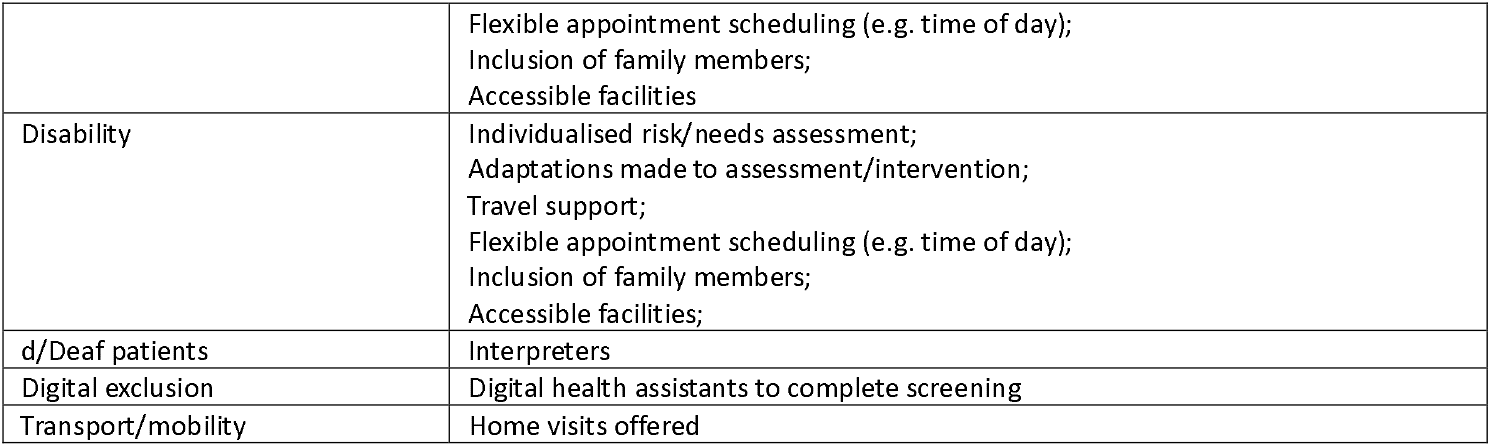
Free-text responses about the tailoring of initial assessments.

Nearly half of respondents (25; 49%) said that they conducted a holistic assessment for patients, while 18 (35%) did not, and six (12%) were unsure. Forty-six (90%) said that they conducted a physical activity assessment, 39 (77%) conducted a nutrition assessment, and 37 (73%) conducted a psychological assessment. Only 11 (22%) said that they conducted a behaviour change assessment.

Here, we highlight the extent to which universal, targeted, and specialist interventions were offered across the physical, nutritional, psychological, and behavioural domains (Supplementary Table 3 includes further detail on the type of interventions provided by prehabilitation services).

For universal interventions, physical interventions (45; 88%) were the most widely implemented, followed by nutritional interventions (42; 82%). Psychological interventions were also provided in most services (41; 81%). Behavioural interventions were delivered in fewer services (27; 53%). ‘Other interventions’ were provided in eight services (16%).

For targeted interventions, physical interventions were delivered in 47 services (92%), nutritional interventions in 39 (76%), psychological interventions in 33 (65%), behavioural interventions in 25 (49%) and other interventions in six (12%).

A similar pattern was observed in specialist prehabilitation in which physical interventions were delivered in 43 services (84%), nutritional interventions in 35 (69%), psychological interventions in 30 (59%), behavioural interventions in 21 (41%), and other interventions in eight services (16%).

Where respondents described ‘other’ interventions in the free text, this referred to smoking cessation, alcohol cessation and fatigue management.

A section of the survey was dedicated to understanding how services addressed health inequalities. Responses are summarised in Table 3.

**Table 3:**
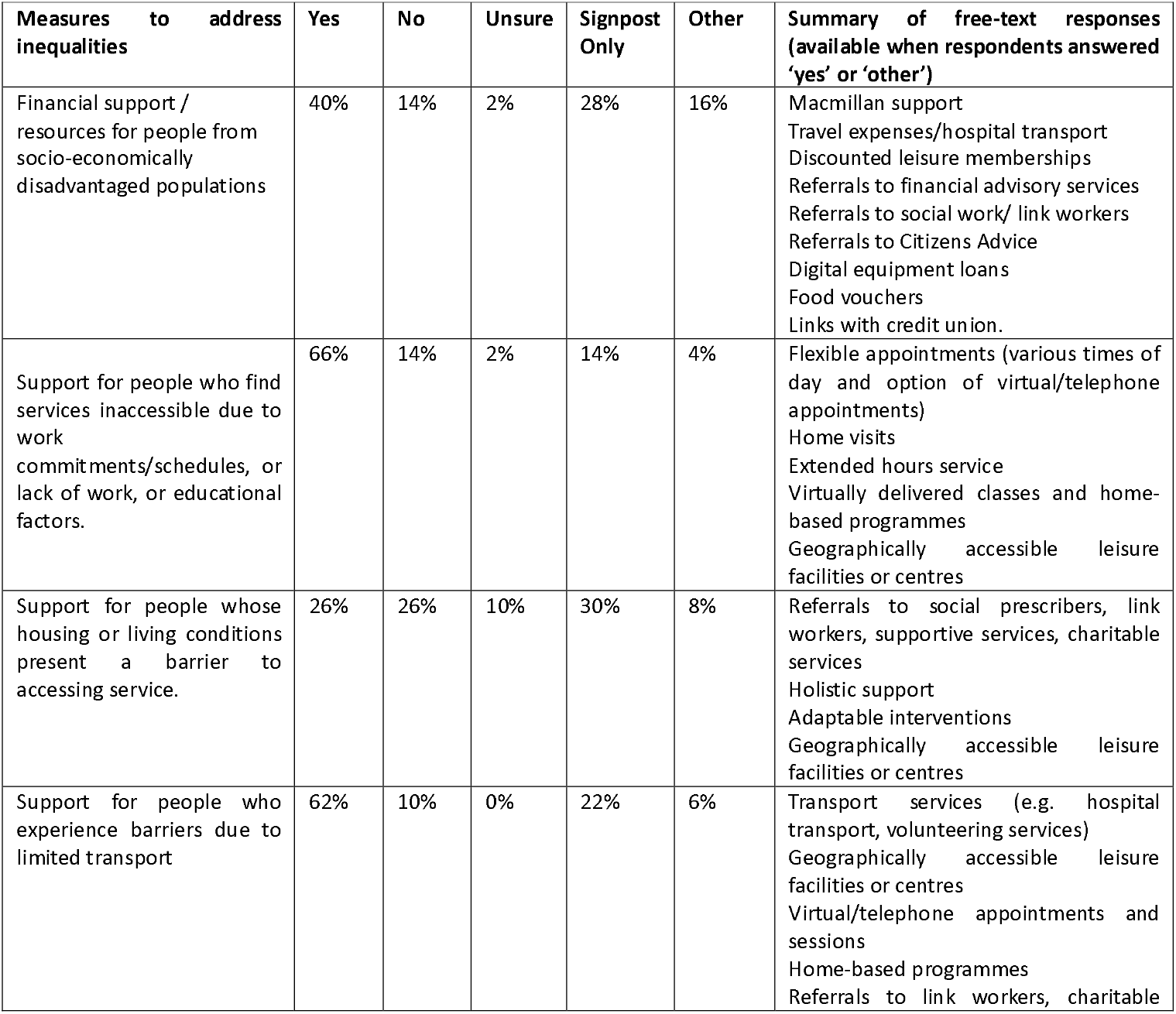
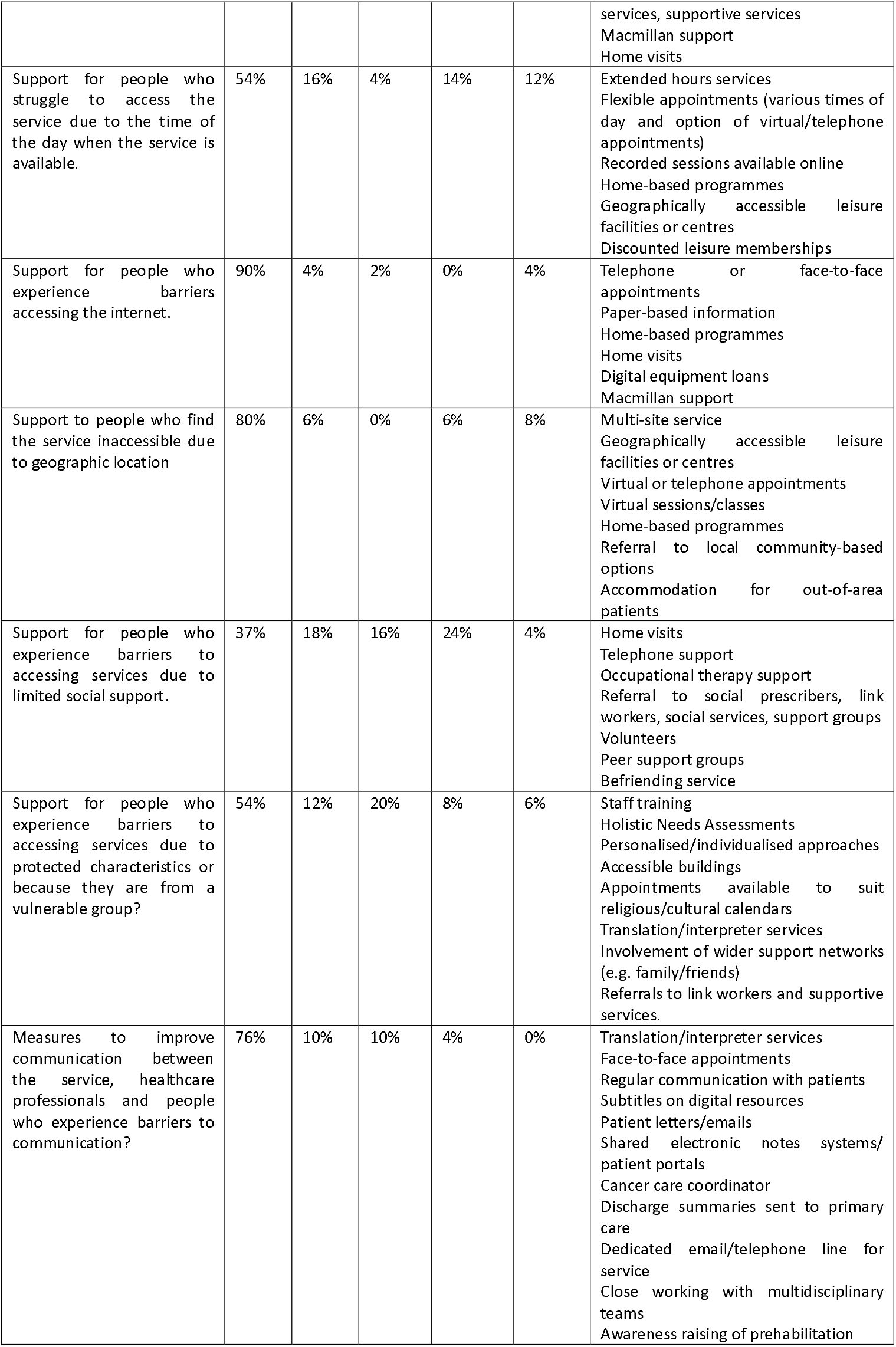
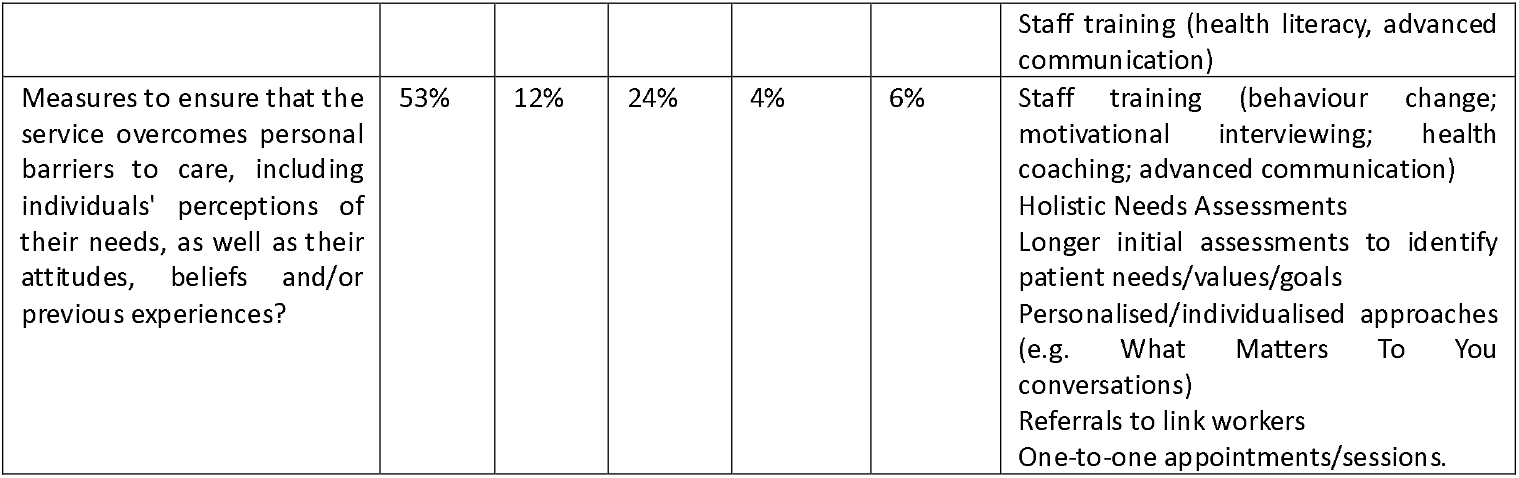
Summary or responses to the ‘Addressing Inequalities’ section.

**Table 4:**
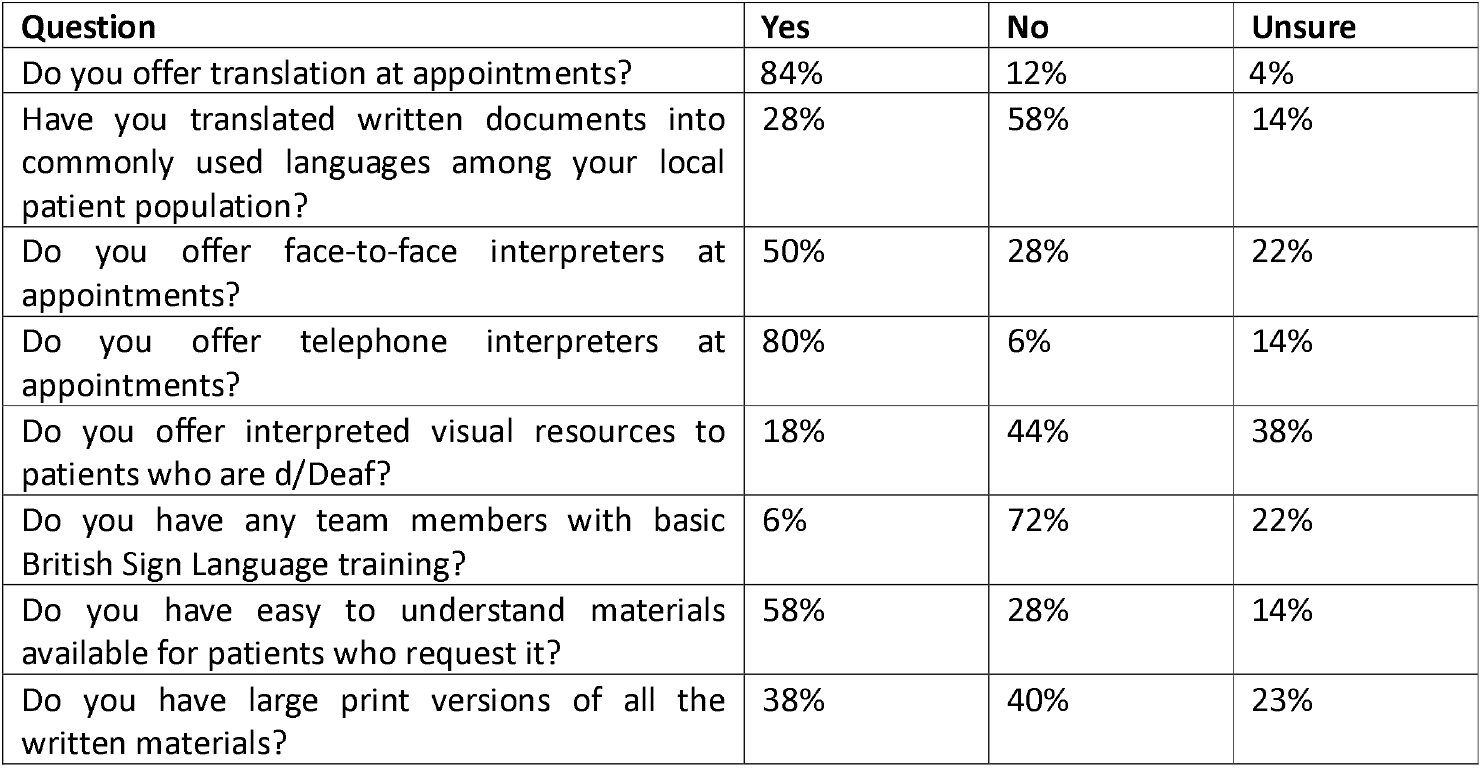
Measures for addressing communication inequalities.

| Question | Yes | No | Unsure |
| --- | --- | --- | --- |
| Do you offer translation at appointments? | 84% | 12% | 4% |
| Have you translated written documents into commonly used languages among your local patient population? | 28% | 58% | 14% |
| Do you offer face-to-face interpreters at appointments? | 50% | 28% | 22% |
| Do you offer telephone interpreters at appointments? | 80% | 6% | 14% |
| Do you offer interpreted visual resources to patients who are d/Deaf? | 18% | 44% | 38% |
| Do you have any team members with basic British Sign Language training? | 6% | 72% | 22% |
| Do you have easy to understand materials available for patients who request it? | 58% | 28% | 14% |
| Do you have large print versions of all the written materials? | 38% | 40% | 23% |

Within the section on addressing inequalities, the survey included questions that focused on addressing communication inequalities, as this was identified as an important topic by our patient and public panel and by patients and healthcare professionals who participated in the Delphi consensus study in an earlier stage of the project [14].

With regards to recording sociodemographic data amongst participating patients to evaluate equity of access to services, respondents indicated that they recorded various sociodemographic characteristics, including: gender (36; 70.6%); ethnicity (29; 57%); sex (29; 57%); socioeconomic status (18; 35%), religion (14; 28%); disability (13; 26%), and language (13; 26%). Other factors (41.2%) were tracked in 21 (41%) services. Where free-text responses were provided, these included: age, indices of multiple deprivation decile, Charlson comorbidity index, home location, political opinion, caring responsibilities, and communication needs.

Regarding research, audit, and quality improvement (RAQI) activities, 27 (53%) services engage in ongoing quality improvement projects, 17 (33%) conduct audits against agreed standards, while seven (14%) have completed independent research studies. Only five respondents (10%) were unaware of any RAQI activities within their service, while two (4%) explicitly stated they do not engage in such activities. Thirty-nine services (77%) indicated that they evaluate their impact, while seven (14%) do not. Four respondents (8%) were unsure, and one (2%) did not respond. Detailed responses to questions regarding the evaluation of services are presented in Supporting Information (Appendix D).

Twelve respondents (24%) were willing to share their evaluation reports with our team. The key outcomes measured in evaluations comprised patient feedback (40; 78.4%), followed by length of hospital stay post-surgery (36; 70.6%), quality of life (27; 53%), peri-operative complications (23; 45%), critical care bed days (23; 45%), and 30-day readmission rates (22; 43%). Less commonly measured metrics included 30-day survival (8; 16%), 1-year survival (6; 12%), and survival to hospital discharge (5; 10%). Twenty-nine services (57%) collected data on other evaluation metrics, which are summarised in Box 3.

### Box 3: additional evaluation measures

EQ5D5L

90-day emergency attendances

90-day readmissions

Visual Fatigue Score

Sit to stand tests

Six-minute walk test

PG-SGA

PHQ-4

EORTC C30

Compliance

DAOH30

Complication Index

Weight loss/gain

Smoking/alcohol status

Handgrip strength

Muscle mass/fat-free mass index

Twenty-four services (47%) tracked patient mortality during the prehabilitation period, while 21 (41%) did not, and five (10%) were unsure. However, after discharge from prehabilitation, mortality tracking was only undertaken in 12 services (24%), with 34 (67%) not tracking it and four respondents (8%) were unsure.

Regarding patient adherence and attendance, non-attendance to screening appointments was recorded by 36 (71%) of services, while 10 (20%) did not track it. Non-attendance at initial assessments was recorded by 43 (84%), making it the most tracked adherence measure. Patient non-adherence to interventions was recorded by 35 services (69%); eight services (16%) did not track it, and six respondents (12%) were unsure.

## Discussion

As far as we are aware, this study represents the first UK-wide mapping exercise of prehabilitation services before cancer surgery, which includes service descriptions and considerations of health inequalities. We are, however, aware of a UK-wide study by Pufulete et al. [19], which obtained responses on prehabilitation services (not specific to cancer surgery) through Freedom of Information requests (for which responses are legally mandated, to provide transparency of information amongst public-sector organisations). A report from Scotland has been published on the provision of prehabilitation for cancer, and a Scotland-wide study undertaken on behalf of the cancer support charity Macmillan [20].

Pufulete *et al*. demonstrated that 78 organisations offered prehabilitation, 22 of which were offered to cancer surgical patients only, and 64 organisations did not offer prehabilitation [19]. These figures are broadly in line with our findings. The Macmillan study of Scottish services showed that stakeholders in cancer care in Scotland appraised prehabilitation as important; however, they noted issues with the clarity of the definition of prehabilitation [20], and some respondents were not aware whether local services were available. Where services were described, they were usually small and primarily focused on the provision of a single discipline-led component [20]. Following the COVID-19 pandemic, many services stopped face-to-face appointments and resumed as hybrid or online-only services, and the number of services with permanent funding decreased [12]. The need for improvements to screening, identification and offers of, and referrals to prehabilitation were identified in both. Two-thirds of respondents did not know if there were plans to introduce or expand prehabilitation activities, and 65% of respondents said that funding was due to end by 2023, with only 2% of services permanently funded. This precarity of funding for prehabilitation corresponds with findings from this survey.

Despite the initial challenges in contacting prehabilitation services, our study ultimately yielded a high response rate from organisations, with 73 organisations indicating that they had a prehabilitation service for patients who were preparing for cancer surgery, and 39 organisations confirming that there was no service. This means that our study obtained at least some information from 96% of eligible NHS organisations. Fifty-one organisations completed the full survey, providing detailed information on 69% of the prehabilitation services that we identified. There was a high level of completion across most survey questions, suggesting strong engagement from participants, most of whom hold leadership roles in prehabilitation services. Most participants shared their professional details and work experience, with only a small percentage of responses missing.

One of the issues that we encountered in multiple aspects of this study is the lack of a clear consensus regarding exactly what prehabilitation is, and therefore how it can be identified and distinguished from other pre-operative services such as pre-anaesthetic assessment clinics. It is clear from our data that a wide range of definitions are in use, and a broad range of practices are considered to fall under the label of prehabilitation. Indeed, there is no single ‘model’ of prehabilitation – both in terms of what is provided, and where the service is located within healthcare organisational structures. This made reaching eligible prehabilitation services difficult but is also suggestive of other challenges within the prehabilitation context. Early guidance suggests that prehabilitation in its first iterations was considered “the first stage in the rehabilitation pathway, otherwise known as preventive rehabilitation” [21], reflecting the focus on preventing treatment-related complications. Since then, prehabilitation has been variously conceptualised as ‘preconditioning’ [22], pre- or peri-operative ‘optimisation’ [23], preparation [24, 25], ‘tertiary prevention’ [26], as well as lifestyle management, and even a form of population health [25]. Considering how these concepts might appeal to different members of the healthcare team, it is easy to see how services might be aligned with different departments and professions, and it is perhaps unsurprising that the two largest groups of respondents in our dataset were physiotherapists (43%) (who have traditionally had a significant role in rehabilitation) and anaesthetists / peri-operative physicians (16%) who have traditionally had a significant role in pre-operative optimisation.

The uncertainty regarding the definition and content of prehabilitation was compounded by the diversity of ‘brand names’ by which programmes were identified, some of which seemed designed to invoke concepts related to exercise, fitness and preparation, but not necessarily healthcare. This is an interesting contrast to most NHS departments, which are simply labelled according to their organisational function or location (e.g., ‘Department of Anaesthesia’, ‘Ward C2’). From our perspective as researchers, this made it challenging to identify and contact services and raises questions about the implications for patients. On the one hand, a ‘brand name’ may be a useful shorthand and could have a role in engaging or motivating patients, but on the other hand, it may make the function of the service less obvious, particularly if patient care moves between organisations. It may also reflect a sense that ‘prehabilitation’ is not always an adequate term to capture and communicate what a service seeks to do [2].

Whilst there may be growing awareness of prehabilitation as a term, there remains uncertainty over definitions and applications, and what exactly constitutes prehabilitation [20]. For example, a literature review on prehabilitation in head and neck cancer found that one-third of the studies defined prehabilitation as *preventative* exercises before the start of acute cancer treatment; the remaining two-thirds defined prehabilitation as *treatment*-*concurrent* prehabilitation [27].

The Macmillan (2017) working model of prehabilitation considered prehabilitation to have three stages [21]:

1. Pre-assessment: to measure the baseline, and identify risk factors amenable to interventions
2. Interventions: to improve the patient’s function; physical activity is “always present”, and dietary and psychological well-being interventions are often present. Other interventions may include smoking cessation, alcohol reduction, respiratory exercises, lymphoedema management, medication and comorbidities review, as well as a raft of other possible interventions which relate to specific patient needs.
3. Follow-up: to determine progress and ensure appropriate follow-up

Within these various articulations of prehabilitation lie several assumptions about the problem that prehab seeks to fix. For example, when conceptualised as ‘prevention’, prehabilitation becomes an intervention to prevent treatment-related complications or mitigate iatrogenic harm. However, ‘optimisation’ approaches position prehabilitation as an opportunity to address ‘health risk behaviours’. As the remit of prehabilitation is widened to include chemotherapy, radiotherapy and other oncological treatments, conceptualisations of ‘what prehabilitation does’ will become more complex (e.g. prehabilitation as improving ‘tolerance’ of, and ‘responsiveness’ to therapies) [28, 29].

We found that the funding of prehabilitation is often unstable, with services tending to be temporarily funded via pilot schemes or patchwork arrangements of nonrecurrent and charitable funding, and with 10% of services unfunded. This is likely to make the financial sustainability and development of services precarious and is itself a potentially important source of inequality (i.e., between areas where prehabilitation is commissioned on a permanent basis, and those where funding is less stable) [6]. One respondent identified funding as a barrier to establishing a formal name for their service, indicating how limited or non-recurrent funding can have an impact on service visibility and identity. This is similarly reported in studies from Scotland, where funding has been identified as a key barrier for services, impacting the ability to attract and retain staff [20]. Precarious resourcing within services is also likely a contributor to broad reliance of services on charity, voluntary and community input, limiting the extent to which responding to inequalities can be embedded at the core of services.

Our findings indicate that physiotherapists and dietitians are the most well-represented professions in services, which may relate to influential publications emphasising the contribution of allied health professionals in prehabilitation [30]. There are fewer psychological professionals involved, although some of the psychological or ‘wellbeing’ offerings of services are likely to be provided by other professions (e.g. wellbeing practitioners). This may, in part, reflect the well-recognised national staffing challenges amongst psychology professionals [31]. Across universal, targeted and specialist models of prehabilitation, physical interventions are the most frequently provided intervention, followed by nutritional, then psychological and behavioural, suggestive of the ‘hierarchy’ of importance and evidence, and in keeping with the early Macmillan model [21]. As we have written elsewhere, this may be because psychological interventions have little evidence of improving commonly measured surgical outcomes [6]. However, given that there are well-known social determinants of mental health [32-34], and that poor mental health may limit a patient’s engagement with healthcare, the relative deficit in formal psychological input may compound inequalities of engagement with prehabilitation services.

Services more frequently provide for those cancers for which there is currently the strongest evidence base (upper and lower gastrointestinal, and lung). However, a relatively large proportion of services provide prehabilitation for breast and gynaecological cancers where evidence is comparatively sparse [35], perhaps suggesting that prehabilitation interventions have been extrapolated as having generalisable benefits [6], or that prehabilitation services are tending to expand to include patients with more common cancer types. Potentially linked to this, we found that research, audit and quality improvement activity was commonplace, and this may be motivated by a desire to generate local evidence of impact in the context of an incomplete evidence base and lack of standardised outcome measures.

Most services provide prehabilitation to fewer than 500 patients each year, indicating that prehabilitation is relatively small-scale when considering that approximately 385,000 people in the UK are diagnosed with cancer every year, and nearly half of these will undergo surgery [36, 37]. This is consistent with the relatively restrictive scope of services (e.g., in terms of eligible cancer types).

Where respondents indicated tailoring to support different characteristics, these were broadly aligned with what might be expected of any high-quality elective healthcare service, for example, translation and interpretation, or accessible facilities [38]. Several services referred to ‘personalisation’ of assessments, however, without detail on how services tailor prehabilitation it is not possible to comment on the quality of this personalisation in meeting different patient needs. This is something that the PARITY team have explored in greater depth in a series of case studies in prehabilitation services in the UK; we will report the findings of that work in due course.

Whilst responses suggest that there are many steps taken to address inequalities within services, the provision of some mitigations is better than others. For example, reference to geographically accessible centres and a virtually accessible service are more commonly reported than embedded responses that support patients for whom financial means or housing is a barrier, which frequently draws on support from voluntary and community sector partners. Similarly, interpretation for languages other than English are more readily supported, whilst there is work to be done on the development of resources for d/Deaf population or groups requiring Easy Read documents.

Overall, we identified that prehabilitation services for patients preparing for cancer surgery were highly variable across the UK, including in terms of eligibility, accessibility, staffing, funding and service design. There is a large body of literature on variation in healthcare which acknowledges that some variation is desirable (e.g., when it exists to respond to the needs of the patients or population served), but it can often be a cause of inequitable healthcare and the lack of consensus about ‘what good looks like’ can make it challenging to uphold high standards [39, 40]. However, while our survey indicates that the services that patients can access are different depending on where they live (and some may arguably be ‘better’ than others), there is no evidence of harm to patients in our data.

Whilst we cannot draw firm conclusions about the reason for variation between services based on the data from this study, there is evidence of multiple possible explanations: prehabilitation is relatively new (with the oldest service being only ten years old at the time of the survey), and so services can be expected to be at various stages of development and expansion; services are also relatively small scale, which may limit the number of staff (and hence, the diversity of staff groups) involved; finally, the challenging funding situation of many prehabilitation services may also be significant as services may be offering what they can for the (limited and time-bound) funds available.

### Limitations and Challenges

Although we believe this is the most comprehensive survey of its type, this paper provides an incomplete picture of the presence of prehabilitation before cancer surgery in the UK. Identifying eligible services was particularly challenging, and where services were identified, some were unable to complete the survey (e.g., due to time constraints). We therefore acknowledge that some services have been missed. Although challenges in recruiting participants are frequently encountered in survey-based research, we suspect that this is of particular significance in prehabilitation due to the relative lack of awareness of services in some organisations. We encountered evidence of this during the study, for example, on several occasions, a representative of an organisation confirmed that there was no eligible service, only for someone else from the same organisation to get in touch to identify their service for participation.

Although the survey sought to capture the different contexts of health services across the four nations of the UK, we acknowledge that the design of the survey reflected the functioning of NHS services in England. Notably, we have not captured the picture of prehabilitation service provision in Scotland due to the eligibility criteria of our protocol (which was based on identifying services that patients receive as part of NHS pathways). This gap in our data is mitigated in part by the existence of a recent national survey of Scottish prehabilitation services by Provan et al [20].

### Recommendations

The landscape of prehabilitation before cancer surgery in the UK is rapidly changing, and this offers an opportunity for services to account for inequalities as they are developed, implemented, evaluated and updated. Our data indicates inequitable access to prehabilitation, which can be addressed on four levels.

Firstly, there remain some areas of the UK where prehabilitation is not available to any patients who are preparing for cancer surgery as part of their NHS care. Based on this, we recommend that health services should take steps to agree on the need for prehabilitation at the national, strategic level, and support individual organisations in implementing services.

Secondly, NHS organisations are inconsistent in the support that they provide to services. Most notably in terms of funding, with most services in receipt of temporary funding and some existing without any funding at all. Whilst we recognise the need to ‘pilot’ new services, this should be done with a view to commissioning services with sustained funding.

Thirdly, there is a need for consensus regarding the definition of prehabilitation, and what it consists of. This created challenges for us in conducting the study and is likely also to create challenges for patients. Related to this, prehabilitation services should be more consistently named, which would make it easier for patients (and researchers) to identify and hence access them.

Finally, services and those who commission them should acknowledge the risk of exacerbating health inequalities through design, implementation and delivery, and take steps to address this. Our previous paper provides consensus guidance on this based on a Delphi process [14], and our subsequent work will expand this based on case studies of actual services.

Furthermore, given the pace of development of prehabilitation, services are likely to change significantly in the coming years; as such, further mapping exercises will be required to maintain knowledge of service provision across the UK.

## Supporting information

Appendix A: PARITY service evaluation survey

Appendix B: Detailed information on the characteristics of prehabilitation services

Appendix C: Detailed information on referral and assessment processes

Appendix D: Detailed responses on the evaluation of services

## Data Availability

All relevant data are within the manuscript and its Supporting Information files.

## Acknowledgements

This work was funded by the National Institute for Health and Care Research (NIHR) Health Services and Delivery Research programme as part of the PARITY study (NIHR134282). The views expressed are those of the authors and not necessarily those of the NIHR or the Department of Health and Social Care. The authors have no conflicts of interest to declare.

^Collaborators: Greg Warren, James Durrand, Stuart Connal, Jon Barnes, Anne Devine, Lorna Starsmore, Sophia Beeby, Neelesh Mohan, Reet Nijjar, Sam Moore, Sarah Peacock.

## Captions for Supporting Information

Appendix A: PARITY service evaluation survey

Appendix B: Detailed information on the characteristics of prehabilitation services

Appendix C: Detailed information on referral and assessment processes

Appendix D: Detailed responses on the evaluation of services

