## Appendix A: PARITY service evaluation survey for "Prehabilitation before cancer surgery in the UK National Health Service: what services exist, and how do they address health inequalities?"

*UK Prehabilitation before Cancer Surgery*
*PARITY Service Evaluation Mapping Questionnaire*

PARITY

**Thank you for participating in this survey.**
**The survey should take between 30-60 minutes to complete.**

Please complete in full with relevant detail for your prehabilitation services in cancer care.

Please read each question and select all answers that apply.

For any questions that do not apply to your current prehabilitation intervention, leave blank '☐' or mark free-text answers as 'N/A'

Please provide additional details describing your service as required.

If you require support, or experience issues with the survey, please contact us via

---

Q1 Are you a TRIPOM associate completing the survey on behalf of a service?

☐Yes
☐No

*If you answered Yes to Q1 please complete Q2*

Q2 If you are a TRIPOM associate completing the survey on behalf of a service please provide your name and email address.

Name: ____________________
Email: ____________________

**Section 1. Prehabilitation in cancer care service characteristics**

For the purpose of this survey, prehabilitation is broadly defined as interventions which are designed to enhance a patient's functional, nutritional and / or psychological capacity and which are delivered following a cancer diagnosis and prior to cancer surgery.

These include interventions that:

- Are part of the funded usual care pathway for patients in the Trust / health board and offered, referred to, or signposted by the NHS cancer care team.
- May be for all cancer types and operations, or specific cancers or operations.
- May be offered universally (all patients), targeted (high risk patients), or specialist (for those with complex needs).
- May be delivered in hospital, community, or online settings, including by a commissioned non- NHS provider (including third sector, commercial and local authorities).

---

**Section 1.1. About your organisation**

Q3 Please state the name of the prehabilitation services provided at your Trust/Hospital

Name (e.g. 'Surgery School'): ______________________________________________

The NHS Trust _________________________________________________________

Hospital _______________________________________________________________

---

Q4 Is your service delivered by your Acute Trust?

☐Yes
☐No
☐Unsure

---

Q5 Please provide the prehabilitation service lead's name and contact email:

Name: ______________________________________________

Email: ______________________________________________

Q6 Are you a prehabilitation service lead?

☐Yes
☐No
☐Unsure

---

Q7 We will develop a national map of prehabilitation services before cancer surgery for improving service delivery.

It would be useful if you could include a generic contact information for your service so that we may include it in the report.

Please also indicate whether you are able to give permission for this information to be shared (Y = yes or N = No).

*Note: This will be publicly available information. Please avoid sharing personal details.*

Email ___________________________________________________________

Telephone _______________________________________________________

Permission to share contact information (Y/N) ______________________________

**Section 1.2. About you**

Q8 What is your role in prehabilitation?

___________________________________________________________

___________________________________________________________

___________________________________________________________

Q9 What is your profession?

Q10 How long have you been employed in this role? (Years, Months)

**Section 2. The size and the characteristics of the prehabilitation service**

Q11 When was the prehabilitation before cancer surgery service first set up at your organisation? (Year only).

*Please provide your best guess if you are unsure.*

Q12 Which department is the prehabilitation service located in? (e.g. physio department, local gym, etc).

Q13 Does your service cover other hospitals in your Trust?

☐Yes

☐No

☐Unsure

☐Not Applicable

---

*If you answered Yes to Q13 please complete Q14*

Q14 Please state the names of all the hospitals that are included in your prehabilitation service **inside your Trust:**

1. ______________________________________________
2. ______________________________________________
3. ______________________________________________
4. ______________________________________________
5. ______________________________________________
6. ______________________________________________
7. ______________________________________________
8. ______________________________________________
9. ______________________________________________
10. ______________________________________________

---

Q15 Does your service cover other hospitals outside of your Trust?

☐Yes

☐No

☐Unsure

☐Not Applicable

---

*If you answered Yes to Q15 please complete Q16*

Q16 Please state the names of all the hospitals that are included in prehabilitation **outside of your Trust?**

1. ____________________________________________

2. ____________________________________________

3. ____________________________________________

4. ____________________________________________

5. ____________________________________________

---

Q17 Does your service provide prehabilitation to patients who live outside your Integrated Care Board area?

☐Yes

☐No

☐Unsure

☐Not Applicable

Q18 How is the service funded?

☐Permanent funding received and implemented

☐Research grant-based service

☐Commissioned service

☐Pilot service with potential to extend

☐Unfunded

☐Other (please describe)

______________________________________________

Q19 If the service is permanently funded, please state who provides funding (optional question).

____________________________________________________________

____________________________________________________________

____________________________________________________________

____________________________________________________________

Q20 Do you undertake research/audit/quality improvement (RAQI) to measure the effectiveness of prehabilitation service before cancer surgery?

*Please select all that apply*

☐No, I'm not aware

☐No, we do not carry out any RAQI activities

☐Yes, there is ongoing funded research

☐Yes, we carried out research and have completed the study

☐Yes, we audit our service against agreed standards

☐Yes, we have on-going quality improvement projects

☐Other (please describe) ____________________________________________

---

Q21 Which of the following specialities/professions participate in your prehabilitation before cancer surgery service?

*Please select all that apply*

☐Physiotherapist

☐Clinical Psychologist

☐Dietitian

☐Health Psychologist

☐Occupational Therapist

☐Nurse

☐Cancer Nurse Specialist

☐Oncologist

☐Anaesthetist

☐Surgeon

Other (please specify) ______________________________________

Other (please specify) ______________________________________

Other (please specify) ______________________________________

---

Q22 Which of the following cancer sites are included in your prehabilitation before cancer surgery service?

*Please select all that apply*

☐Brain and Central Nervous System

☐Head and Neck

☐Gynaecological

☐Upper Gastrointestinal Tract

☐Lower Gastrointestinal Tract

☐Sarcomas

☐Lung

☐Haematological

☐Breast

☐Urological

☐Skin

☐Non-site specific

☐Childhood

☐Liver and Pancreas

Other (please specify) ________________________________________________

---

Q23 On average, how many cancer patients per year do you see in your service?

*Please provide an estimate number based on your experience*

☐Fewer than 500 patients

☐501-999 patients

☐More than 1000 patients

---

Q24 Is your prehabilitation service also offered to oncology only patients? (e.g. chemotherapy and radiotherapy)

☐Yes

☐No

☐Unsure

---

Q25 Is your prehabilitation service offered to palliative care patients?

☐Yes

☐No

☐Unsure

---

Q26 Is your service offered to other patient groups outside of cancer?

☐Yes

☐No

☐Unsure

---

Q27 Do you have formal agreements with any community-based, third-party or independent providers outside the hospital settings?

☐Yes

☐No

☐Unsure

---

*If you answered Yes to Q27 please complete the following table (Q28)*

Q28 If you have formal agreements with any community-based, third-party or independent providers, please state further details below.

| **Organisation Name** (e.g. gym) | **Type of prehabilitation provided** (e.g. exercise class) | **By whom** (e.g. physiotherapist with cancer speciality) | **How they are used** (e.g. patients are signposted for self-referral) |
|---|---|---|---|

Q29 Do you have any **informal** agreements with community-based, third-party or independent providers outside the hospital settings?

☐Yes

☐No

☐Unsure

*If you answered Yes to Q29 please give more details here (Q30)*

Q30 If yes, what are the most commonly used informal providers that patients are signposted to? (e.g. Macmillan walks)

1. ______________________________________________

2. ______________________________________________

3. ______________________________________________

4 ______________________________________________

**Section 3. Screening and individualised assessment for prehabilitation**

Macmillan principles and guidance for prehabilitation states that identification of people with cancer who require prehabilitation (i.e. screening) should occur as early as possible from diagnosis (and in some cases before a confirmed diagnosis), and in advance of each treatment.

Screening should use validated tools to identify the need for more detailed assessment in order to inform the prescription of targeted or specialist interventions.

Screening should be aligned to the Holistic Needs Assessment (HNA) and should include psychological risk factors, physical fitness and nutrition including evaluation of weight loss, poor intake, body mass index and nutrition impact symptoms.

Individualised assessment, when indicated, should encompass comprehensive evaluation of needs identified during screening using validated clinical measurement techniques. Assessments should inform the individualised prescription of exercise, nutrition and psychological interventions.

---

Q31 Do you have a defined referral criterion to prehabilitation before cancer surgery in your service?

☐Yes

☐No

☐Unsure

Q32 When are referrals to screening and initial individualised assessments carried out?

☐At the same time

☐At different appointments

☐Unsure

Q33 Who refers patients to the screening assessment for prehabilitation before cancer surgery?

*Please select all that apply*

☐General Practitioner

☐Nurse

☐Cancer Nurse Specialist

☐Anaesthetist

☐Self-referral

☐Oncologist

☐Surgery

Other (please describe) ________________________________

---

Q34 How does the person who carries out referral communicate prehabilitation with patients?

*Please select all that apply*

☐Phone

☐Letter

☐Mobile Phone portal/apps

☐Electronic

☐Face-to-face

☐Other (please describe) ____________________________________________

Q35 Where does the initial assessment appointment take place?

*Please select all that apply*

☐Primary care

☐Hospital

☐Home-based

☐Community centre

☐Telephone based

Other (please describe) _______________________________________________

---

Q36 Who carries out the initial assessment?

*Please select all that apply*

☐Nurse

☐Cancer Specialist Nurse

☐Prehabilitation Coordinator

☐Physiotherapist

☐Dietitian

☐Psychologist

☐Surgeon

☐Anaesthetist

☐Occupational Therapist

☐Administration Assistant

Other (please describe) ____________________________________________________

Q37 When is the initial assessment carried out?

☐Shortly after cancer diagnosis
☐Shortly after the decision for surgery
☐Before surgery during pre-operative assessments
☐All of the above
☐No specified time
☐I don't know

Q38 Is the initial assessment tailored to different groups of patients? (e.g. by sex, language etc).

☐Yes
☐No

*If you answered Yes to Q38 please complete the table below (Q39) with further details*

Q39 Please give further detail on how initial assessments are tailored to different groups

| | How do you tailor the assessment to different groups?<br><br>Please give details below. If not applicable, please use 'N/A' |
|---|---|
| Sex (e.g. Male only, female only) | |
| Language | |
| Cultural, ethnic and religious differences | |
| Learning disability | |
| Family orientation | |
| Other | |
| Other | |
| Other | |

In the next section we would like to ask you about the tools you use during initial assessment.

This will include assessment measures for:

- Holistic Assessment
- Physical Activity Assessment
- Nutrition Assessment
- Psychological Assessment
- Behaviour Change Assessment

Please state the names of the measures you use for each question, and/or scales.

Q40 Do you conduct a holistic assessment during initial assessment?

☐Yes

☐No

☐Unsure

*If you answered Yes to Q40 please complete Q41 below*

Q41 What are the tools that are being used during **holistic assessment**?

Please state the names of the measures used, and/or scales.

| **Tools used**<br>e.g Holistic Needs Assessment | **Is this a standardised measure?**<br><br>**Yes/No/Adapted/Unsure** |
|---|---|

Q42 Do you conduct a physical activity assessment during initial assessment?

☐Yes
☐No
☐Unsure

*If you answered Yes to Q42 please complete Q43 below*

Q43 What are the tools that are being used during **physical activity assessment**?

Please state the names of the measures used, and/or scales.

| **Tools used**<br>e.g Cardio Pulmonary Exercise Testing (CPET) | **Is this a standardised measure?**<br><br>**Yes/No/Adapted/Unsure** |
|---|---|

Q44 Do you conduct a nutrition assessment during initial assessment?

☐Yes

☐No

☐Unsure

*If you answered Yes to Q44 please complete Q45*

Q45 What are the tools that are being used during **nutrition assessment?**

Please state the names of the measures used, and/or scales.

| **Tools used**<br>e.g Body Mass Index | **Is this a standardised measure?**<br><br>**Yes/No/Adapted/Unsure** |
|---|---|

Q46 Do you conduct a psychological assessment during initial assessment?

- Yes
- No
- Unsure

*If you answered Yes to Q46 please complete Q47*

Q47 What are the tools that are being used during **psychological assessment?**

Please state the names of the measures used, and/or scales.

| **Tools used**<br>e.g Hospital Anxiety and Depression Scale (HADS) | **Is this a standardised measure?**<br><br>**Yes/No/Adapted/Unsure** |
|---|---|

Q48 Do you conduct a behaviour change assessment during initial assessment?

- Yes
- No
- Unsure

*If you answered Yes to Q48 please complete Q49*

Q49 What are the tools that are being used during **behaviour change assessment?**

Please state the names of the measures used, and/or scales.

| **Tools used**<br>e.g Patient Activation Measure (PAM) | **Is this a standardised measure?**<br><br>**Yes/No/Adapted/Unsure** |
|---|---|

**Section 4. Interventions**

**Universal Interventions**

According to Macmillan principles and guidance for Prehabilitation, **universal interventions** are applicable to anyone with cancer.

People with cancer and their families, should receive dietary, exercise and psychological advice and behaviour change support, be sign-posted to appropriate resources, and be advised on how to self-manage, recognise and respond to any change in physical and/or psychological state.

These could include conversations as part of personalised care, providing leaflets, tips and signposting tools and resources that are available (e.g. One You, Moving Medicine etc.)

---

Q50 Which of the following **universal interventions** are being provided in your service? (Please select all that apply)

| | Yes | No | Unsure |
|---|---|---|---|
| Physical | ☐ | ☐ | ☐ |
| Nutrition | ☐ | ☐ | ☐ |
| Psychological | ☐ | ☐ | ☐ |
| Behavioural | ☐ | ☐ | ☐ |
| Other (please describe) | ☐ | ☐ | ☐ |

---

**Targeted Interventions**

According to Macmillan principles and guidance for Prehabilitation, **targeted interventions** are applicable to those people with cancer with and at risk of late effects of disease or treatment and those with other long-term conditions.

Specific needs identified during screening should be addressed with prescribed exercise, nutrition and psychological interventions and behaviour change support by a registered health and care professional according to need.

Adherence and effectiveness should be monitored. Patients who receive these interventions need support based on their disease, treatment, side effects and co-morbidities.

---

Q51 Which of the following **targeted interventions** are being provided in your service? (Please select all that apply)

| | Yes | No | Unsure |
|---|---|---|---|
| Physical | ☐ | ☐ | ☐ |
| Nutrition | ☐ | ☐ | ☐ |
| Psychological | ☐ | ☐ | ☐ |
| Behavioural | ☐ | ☐ | ☐ |
| Other (please describe) | ☐ | ☐ | ☐ |

---

**Specialist Interventions**

According to Macmillan principles and guidance for Prehabilitation, **specialist interventions** are applicable to people with cancer who have complex needs or complex treatment (e.g. major surgery, severe impairment and/or disability) and **will need referral to registered professionals to prescribe exercise, nutrition and psychological interventions** and behaviour change support according to need.

---

Q52 Which of the following **specialist interventions** are being provided in your service? (Please select all that apply)

| | Yes | No | Unsure |
|---|---|---|---|
| Physical | ☐ | ☐ | ☐ |
| Nutrition | ☐ | ☐ | ☐ |
| Psychological | ☐ | ☐ | ☐ |
| Behavioural | ☐ | ☐ | ☐ |
| Other (please describe) | ☐ | ☐ | ☐ |

**Section 5. Addressing Inequalities**

In the following section we would like to ask you about the support you provide to patients to address health inequalities when accessing prehabilitation services.

In the context of cancer services, health inequalities can be seen in the differences between individuals' cancer experience or outcome which result from their social-economic status, race, age, gender, disability, religion or belief, sexual orientation, cancer type or geographical location. Improving access to services involves helping people to command appropriate health care resources in order to preserve or improve their health.

---

Q53 Do you offer financial support/resources to improve access to your service, particularly to those from socio-economically disadvantaged populations?

☐Yes (please describe)

☐No

☐Unsure

☐Signposting only

☐Other (please describe)

---

Q54 Do you offer any support to people who find services inaccessible due to work commitments/schedules, lack of work and/or education?

☐Yes (please describe)

☐No

☐Unsure

☐Signposting only

☐Other (please describe)

Q55 Do you offer any support for people whose housing or living conditions is a barrier to accessing services?

☐Yes (please describe)

☐No
☐Unsure

☐Signposting only

☐Other (please describe)

---

Q56 Do you offer any support to people who may experience barriers due to limited transport?

☐Yes (please describe)

☐No
☐Unsure

☐Signposting only

☐Other (please describe)

---

Q57 Do you offer any support for people who may struggle to access the service due to the time of the day when the service is available?

☐Yes (please describe)

☐No
☐Unsure

☐Signposting only

☐Other (please describe)

Q58 Do you offer any support for people who may experience barriers accessing the internet?

☐Yes (please describe)

☐No
☐Unsure

☐Signposting only

☐Other (please describe)

---

Q59 Do you offer any support to people who find the service inaccessible due to geographic location?

☐Yes (please describe)

☐No
☐Unsure

☐Signposting only

☐Other (please describe)

---

Q60 Do you offer any support for people who may experience barriers to accessing services due to limited social support?

☐Yes (please describe)

☐No
☐Unsure

☐Signposting only

☐Other (please describe)

---

Q61 Do you offer any support to people who may experience barriers to accessing services due to their protected characteristics or because they are from a vulnerable group? E.g. age, disability, gender reassignment, marriage and civil partnership, pregnancy and maternity, race, religion or belief, gender, sexual orientation or people who are socio-economically disadvantaged.

☐Yes (please describe)

☐No
☐Unsure

☐Signposting only

☐Other (please describe)

---

Q62 Does your service have anything in place to improve communication between the service, healthcare professionals and recipients of care, particularly for people who experience barriers to communication?

☐Yes (please describe)

☐No
☐Unsure

☐Signposting only

☐Other (please describe)

---

Q63 Do you have anything in place to ensure that the service overcomes personal barriers to care, including individuals' perceptions of their needs, as well as their attitudes, beliefs and/or previous experiences?

☐Yes (please describe)

☐No
☐Unsure

☐Signposting only

☐Other (please describe)

---

Q64 Do you offer translation at appointments?

☐Yes
☐No
☐Unsure

---

Q65 Have you translated written documents to commonly used languages among your local patient population?

☐Yes
☐No
☐Unsure

---

Q66 Do you offer face-to-face interpreters at appointments?

☐Yes
☐No
☐Unsure

---

Q67 Do you offer telephone interpreters at appointments?

☐Yes
☐No
☐Unsure

---

Q68 Do you offer interpreted visual resources to patients who are d/Deaf?

☐Yes
☐No
☐Unsure

---

Q69 Do you have any team members with basic British Sign Language training?

☐Yes
☐No

☐Unsure

---

Q70 Do you have easy to understand materials available for patients who request it?

☐Yes
☐No

☐Unsure

---

Q71 Do you have accessible large printed versions of all the written materials?

☐Yes
☐No

☐Unsure

---

Q72 Can patients access information about their Personalised Prehabilitation Care Plan (PPCP) 24/7?

☐Yes
☐No

☐Unsure

☐I don't know what it is

*If you answered No or Unsure for Q72 please complete Q73*

Q73 If you answered no or unsure, what is the current accessibility of care plans for the patients?

*E.g. PPCPs can be printed and posted upon request from patients but not available in digital format*

Q74 Are patients and family members able to contact someone who is part of their prehabilitation care team?

☐Yes
☐No
☐Unsure

---

Q75 Can patients' family members, carers and others attend appointments?

☐Yes
☐No
☐Unsure

**Section 6. Evaluation of the services**

Q76 Do you carry out **formal evaluation** of the prehabilitation services?

☐Yes
☐No
☐Unsure

*If you answered No or Unsure to Q76 please skip to Q80*

*If you answered Yes to Q76 please proceed to Q77*

Q77 When was the service last evaluated? (year)

________________________________________________________

---

Q78 Is it possible to share the evaluation?

☐Yes
☐No
☐Unsure

---

*If you answered Yes to Q78 please indicate if you are happy to send by email or state other method*

Q79 If yes, can you send an accessible link or upload documents to us by

________________________________________________________

---

Q80 What is the primary evidence/guidelines that informed the design and delivery of your services? *e.g. a randomised controlled trial, benchmarking from the other services.*

Please state them below:

______________________________________________________________________

______________________________________________________________________

______________________________________________________________________

______________________________________________________________________

______________________________________________________________________

Q81 What are the key outcomes measured in your service/evaluation?

*Please select all that apply*

☐Length of hospital stay post-surgery

☐Amount of critical care bed days

☐Quality of Life (e.g. SF-36)

☐30-day survival

☐30-day readmission

☐Complications

☐Survival to Hospital Discharge

☐1-year survival

☐Patient feedback (PROMS, PREMS)

☐Other (please describe) _______________________________________________

Q82 Do you record and evaluate patient mortality during their time under the prehabilitation service?

☐Yes
☐No

☐Unsure

Q83 Do you record and evaluate patient mortality after discharge from prehabilitation service?

☐Yes
☐No
☐Unsure

---

Q84 Do you record non-attendance to screening appointments?

☐Yes
☐No
☐Unsure

---

Q85 Do you record non-attendance to initial assessment appointments?

☐Yes
☐No
☐Unsure

---

Q86 Do you record patient non-adherence to interventions?

☐Yes
☐No
☐Unsure

---

Q87 Do you evaluate changes in patients' quality of life outcomes pre- and post- prehab?

☐Yes
☐No
☐Unsure

---

Q88 Do you record and evaluate readmission rates?

☐Yes
☐No
☐Unsure

---

Q89 Which of the patient sociodemographic characteristics below are recorded and used for evaluating potential inequalities in care?

*Please select all that apply*

☐Ethnicity
☐Gender
☐Sex
☐Socioeconomic Status
☐Disability
☐Language
☐Religion
☐Other (please describe) ________________________________
☐Other (please describe) ________________________________
☐Other (please describe) ________________________________

**Section 7. Key Examples (optional)**

We would like to collate examples of prehabilitation service delivery to understand similarities and differences in implementation across the United Kingdom.

These examples will be used to identify the barriers and facilitators in implementation of prehabilitation and inform the development of best practice guidelines aiming to minimise inequalities arising from the variation in implementation.

**Intervention 1:** *Please describe an element of your prehabilitation service that has been implemented most successfully in your service, and you would consider to be effective.*

**Intervention 2:** *Please describe an element of your prehabilitation service that you have struggled to implement, or could be improved.*

---

Q90 **Intervention 1**:

Please describe an element of your prehabilitation service **that has been implemented most successfully in your service, and you would consider to be effective.**

- ○ Please state type of the intervention (e.g. Universal nutrition intervention, specialist physical intervention, targeted psychological intervention etc) ________________________________________________
- ○ What does it involve? ________________________________________________
- ○ What is the main aim? ________________________________________________
- ○ Who delivers it? ________________________________________________
- ○ Why is it the most effective intervention in your service? ________________________________________________

---

Q91 **Intervention 2**:

Please describe an element of your prehabilitation service **that has been implemented but could be improved in your service**

○ Please state type of the intervention (e.g. Universal nutrition intervention, specialist physical intervention, targeted psychological intervention etc) ________________________________________________

○ What does it involve? ________________________________________________

○ What is the main aim? ________________________________________________

○ Who delivers it? ________________________________________________

○ Why does this intervention require improvements? ________________________________________________

**Thank you very much for completing the PARITY Prehabilitation mapping questionnaire.**

We will use information from surveys to identify eligible prehabilitation services for participation in case studies.

If your service is eligible, a member of the research team will be in touch to provide more information and to organise next steps.

You can also follow the project on Twitter: @PARITYStudy
and on Facebook: PARITY Study
