## Appendix B: Detailed information on the characteristics of prehabilitation services for "Prehabilitation before cancer surgery in the UK National Health Service: what services exist, and how do they address health inequalities?"

| **Survey questions (main)** | **Sub questions** | **Frequency (N=51)** | **Percentage (%)** | **Summary Findings** |
| --- | --- | --- | --- | --- |
| Please state the name of the prehabilitation services provided at your Trust/Hospital | The name of Trusts | 49 | 96.1 | Summary of names |
|  | The name of Hospitals | 48 | 94.1 |  |
|  | The name of the prehabilitation services | 46 | 90.2 |  |
| Is your service delivered by your Acute Trust? | Yes | 41 | 80.4 |  |
|  | No | 6 | 11.8 |  |
|  | Unsure | 2 | 3.9 |  |
|  | No answer | 2 | 3.9 |  |
| When was the prehabilitation before cancer surgery service first set up at your organisation? (Year only). | Answered (earliest 2014, one hasn’t open yet) | 48 | 94.1 | There are 41 valid year entries, within these giving years:  Range=13 Median=2021  IQR=4years |
|  | No answer | 3 | 5.9 |  |
| Which department is the prehabilitation service located in? (e.g. physio department, local gym, etc). | Answered (varieties) | 48 | 94.1 | Major departments include Physiotherapy dept (17), Cancer service (7), Therapy dept (6), and Leisure Centre (5). |
|  | No answer | 2 | 3.9 |  |
| Does your service cover other hospitals in your Trust? | Yes | 26 | 51.0 |  |
|  | No | 14 | 27.5 |  |
|  | Miss data | 11 | 21.6 |  |
| Does your service provide prehabilitation to patients who live outside your Integrated Care Board area? | Yes | 21 | 41.2 |  |
|  | No | 14 | 43.1 |  |
|  | No answer | 11 | 11.8 |  |
|  | Unsure | 2 | 3.9 |  |
| How is the service funded? | Permanent funding received and implemented | 19 | 37.3 |  |
|  | Research grant-based service | 0 | 0.0 |  |
|  | Commissioned service | 3 | 5.9 |  |
|  | Pilot service with potential to extend | 23 | 45.1 |  |
|  | Unfunded | 5 | 9.8 |  |
|  | Other (please describe) | 18 | 35.3 | Including funding from Prehabilitation Project Lead role funding, Cancer Alliance, Macmillan project managers funds, Cancer Research, Local Authorities/Councils, In-house Charitable Funds, Leisure Centres subsidize, Charity, and Cancer Association. |
| Which of the following specialities/professions participate in your prehabilitation before cancer surgery service? | Physiotherapist | 43 | 84.3 |  |
|  | Dietitian | 37 | 72.5 |  |
|  | Other (please specify) | 33 | 64.7 |  |
|  | Cancer Nurse Specialist | 21 | 42.2 |  |
|  | Anaesthetist | 19 | 37.3 |  |
|  | Occupational Therapist | 17 | 33.3 |  |
|  | Surgeon | 14 | 27.5 |  |
|  | Clinical Psychologist | 11 | 21.6 |  |
|  | Nurse | 8 | 15.7 |  |
|  | Oncologist | 6 | 11.8 |  |
|  | Health Psychologist | 3 | 5.9 |  |
| Which of the following cancer sites are included in your prehabilitation before cancer surgery service? | Lower Gastrointestinal Tract | 37 | 72.5 |  |
|  | Lung | 25 | 49.0 |  |
|  | Upper Gastrointestinal Tract | 22 | 43.1 |  |
|  | Gynaecological | 21 | 41.2 |  |
|  | Urological | 20 | 39.2 |  |
|  | Other (please specify) | 20 | 39.2 |  |
|  | Liver and Pancreas | 16 | 31.4 |  |
|  | Head and Neck | 14 | 27.5 |  |
|  | Breast | 11 | 21.6 |  |
|  | Haematological | 10 | 19.6 |  |
|  | Sarcomas | 5 | 9.8 |  |
|  | Skin | 5 | 9.8 |  |
|  | Brain and Central | 3 | 5.9 |  |
|  | Non-site specific | 2 | 3.9 |  |
|  | Childhood | 0 | 0.0 |  |
| On average, how many cancer patients per year do you see in your service? | Fewer than 500 patients | 36 | 70.6 |  |
|  | 501-999 patients | 9 | 17.6 |  |
|  | More than 1000 patients | 2 | 3.9 |  |
|  | No answer | 4 | 7.8 |  |
| Is your prehabilitation service also offered to oncology only patients? (e.g. chemotherapy and radiotherapy) | Yes | 32 | 62.7 |  |
|  | No | 15 | 29.4 |  |
|  | Unsure | 1 | 2.0 |  |
|  | No answer | 3 | 5.9 |  |
| Is your prehabilitation service offered to palliative care patients? | Yes | 22 | 43.1 |  |
|  | No | 23 | 45.1 |  |
|  | Unsure | 3 | 5.9 |  |
|  | No answer | 3 | 5.9 |  |
| Is your service offered to other patient groups outside of cancer? | Yes | 14 | 27.5 |  |
|  | No | 34 | 66.7 |  |
|  | Unsure | 1 | 2.0 |  |
|  | No answer | 2 | 3.9 |  |
| Do you have formal agreements with any community-based, third-party or independent providers outside the hospital settings? | Yes | 17 | 33.3 |  |
|  | No | 31 | 60.8 |  |
|  | Unsure | 1 | 2.0 |  |
|  | No answer | 2 | 3.9 |  |
| If you have formal agreements with any community-based, third-party or independent providers, please state further details below. | Type of prehabilitation provided  (e.g. exercise class) | 34 | 66.7 | The most exercise classes conducted at local leisure centres. Few other sessions conducted at local councils or universities. |
|  | By whom  (e.g. physiotherapist with cancer speciality) | 37 | 72.5 | Most led by a physiotherapist or a PT instructor. |
|  | How they are used (e.g. patients are signposted for self-referral) | 43 | 84.3 |  |
| Do you have any informal agreements with community-based, third-party or independent providers outside the hospital settings? | Yes | 23 | 45.1 |  |
|  | No | 24 | 47.1 |  |
|  | Unsure | 2 | 3.9 |  |
|  | No answer | 2 | 3.9 |  |
