## Appendix C: Detailed information on referral and assessment processes for "Prehabilitation before cancer surgery in the UK National Health Service: what services exist, and how do they address health inequalities?"

| **Survey questions (main)** | **Sub questions** | **Frequency (N=51)** | **Percentage (%)** |
| --- | --- | --- | --- |
| Do you have a defined referral criterion to prehabilitation before cancer surgery in your service? | Yes | 43 | 84.3 |
|  | No | 7 | 13.7 |
|  | No answer | 1 | 2.0 |
|  | Unsure | 0 | 0.0 |
| When are referrals to screening and initial individualised assessments carried out? | At the same time | 21 | 41.2 |
|  | At different appointments | 24 | 47.1 |
|  | Unsure | 5 | 9.8 |
|  | No answer | 1 | 2.0 |
| Who refers patients to the screening assessment for prehabilitation before cancer surgery? Please select all that apply | Cancer Nurse Specialist | 46 | 90.2 |
|  | Surgery | 27 | 52.9 |
|  | Anaesthetist | 21 | 41.2 |
|  | Oncologist | 20 | 39.2 |
|  | Other (please describe) | 15 | 29.4 |
|  | Nurse | 6 | 11.8 |
|  | Self-referral | 3 | 5.9 |
|  | General Practitioner | 0 | 0.0 |
| How does the person who carries out referral communicate prehabilitation with patients? Please select all that apply | Phone | 28 | 54.9 |
|  | Letter | 5 | 9.8 |
|  | Mobile Phone portal/apps | 3 | 5.9 |
|  | Electronic | 10 | 19.6 |
|  | Face-to-face | 42 | 82.4 |
|  | Other (please describe) | 8 | 15.7 |
| Where does the initial assessment appointment take place? Please select all that apply | Primary care | 0 | 0.0 |
|  | Hospital | 31 | 60.8 |
|  | Home-based | 4 | 7.8 |
|  | Community centre | 13 | 25.5 |
|  | Telephone based | 27 | 52.9 |
|  | Other (please describe) | 12 | 23.5 |
| Who carries out the initial assessment? Please select all that apply | Physiotherapist | 41 | 80.4 |
|  | Other | 22 | 43.1 |
|  | Dietitian | 16 | 31.4 |
|  | Occupational Therapist | 9 | 17.6 |
|  | Cancer Specialist Nurse | 9 | 17.6 |
|  | Prehabilitation Coordinator | 6 | 11.8 |
|  | Anaesthetist | 4 | 7.8 |
|  | Administration Assistant | 1 | 2.0 |
|  | Surgeon | 1 | 2.0 |
|  | Nurse | 1 | 2.0 |
|  | Psychologist | 0 | 0.0 |
| When is the initial assessment carried out? | Shortly after cancer diagnosis | 10 | 19.6 |
|  | Shortly after the  decision for surgery | 11 | 21.6 |
|  | Before surgery  during pre-operative assessments | 1 | 2.0 |
|  | All of the above | 25 | 49.0 |
|  | No specified time | 3 | 5.9 |
|  | I don't know | 0 | 0.0 |
|  | No answer | 1 | 2.0 |
| Is the initial assessment tailored to different groups of patients? (e.g. by sex, language etc). | Yes | 24 | 47.0 |
|  | No | 26 | 51.0 |
|  | No answer | 1 | 2.0 |
| Do you conduct a holistic assessment during initial assessment? | Yes | 25 | 49.0 |
|  | No | 18 | 35.3 |
|  | Unsure | 6 | 11.8 |
|  | No answer | 2 | 3.9 |
| Do you conduct a physical activity assessment during initial assessment? | Yes | 46 | 90.2 |
|  | No | 3 | 5.9 |
|  | Unsure | 0 | 0.0 |
|  | No answer | 2 | 3.9 |
| Do you conduct a nutrition assessment during initial assessment? | Yes | 39 | 76.5 |
|  | No | 10 | 19.6 |
|  | Unsure | 0 | 0.0 |
|  | No answer | 2 | 3.9 |
| Do you conduct a psychological assessment during initial assessment? | Yes | 37 | 72.5 |
|  | No | 12 | 23.5 |
|  | No answer | 2 | 3.9 |
| Do you conduct a behaviour change assessment during initial assessment? | Yes | 11 | 21.6 |
|  | No | 34 | 66.7 |
|  | Unsure | 4 | 7.8 |
|  | No answer | 2 | 3.9 |
