## Appendix D: Detailed responses on the evaluation of services for "Prehabilitation before cancer surgery in the UK National Health Service: what services exist, and how do they address health inequalities?"

**Appendix D: Detailed information on the evaluation of services**

| **Survey questions (main)** | **Sub questions** | **Frequency (N=51)** | **Percentage (%)** |
| --- | --- | --- | --- |
| Do you undertake research/audit/quality improvement (RAQI) to measure the effectiveness of prehabilitation service before cancer surgery? | No, I'm not aware | 5 | 9.8 |
|  | No, we do not carry out any RAQI activities | 2 | 3.9 |
|  | Yes, there is ongoing funded research | 4 | 7.8 |
|  | Yes, we carried out research and have completed the study | 7 | 13.7 |
|  | Yes, we audit our service against agreed standards | 17 | 33.3 |
|  | Yes, we have on-going quality improvement projects | 27 | 52.9 |
|  | Other (please describe) | 14 | 27.5 |
| Do you carry out formal evaluation of the prehabilitation services? | Yes | 39 | 76.5 |
|  | No | 7 | 13.7 |
|  | Unsure | 4 | 7.8 |
|  | No answer | 1 | 2.0 |
| What are the key outcomes measured in your service/evaluation? Please select all that apply | Length of hospital stay post-surgery | 36 | 70.6 |
|  | Amount of critical care bed days | 23 | 45.1 |
|  | Quality of Life (e.g. SF-36) | 27 | 52.9 |
|  | 30-day survival | 8 | 15.7 |
|  | 30-day readmission | 22 | 43.1 |
|  | Complications | 23 | 45.1 |
|  | Survival to Hospital Discharge | 5 | 9.8 |
|  | 1-year survival | 6 | 11.8 |
|  | Patient feedback | 40 | 78.4 |
|  | Other | 29 | 56.9 |
| Do you record and evaluate patient mortality during their time under the prehabilitation service? | Yes | 24 | 47.1 |
|  | No | 21 | 41.2 |
|  | Unsure | 5 | 9.8 |
|  | No answer | 1 | 2.0 |
| Do you record and evaluate patient mortality after discharge from prehabilitation service? | Yes | 12 | 23.5 |
|  | No | 34 | 66.7 |
|  | Unsure | 4 | 7.8 |
|  | No answer | 1 | 2.0 |
| Do you record non-attendance to screening appointments? | Yes | 36 | 70.6 |
|  | No | 10 | 19.6 |
|  | Unsure | 3 | 5.9 |
|  | No answer | 2 | 3.9 |
| Do you record non-attendance to initial assessment appointments? | Yes | 43 | 84.3 |
|  | No | 5 | 9.8 |
|  | Unsure | 1 | 2.0 |
|  | No answer | 2 | 3.9 |
| Do you record patient non-adherence to interventions? | Yes | 35 | 68.6 |
|  | No | 8 | 15.7 |
|  | Unsure | 6 | 11.8 |
|  | No answer | 2 | 3.9 |
| Do you evaluate changes in patients’ quality of life outcomes pre- and post- prehab? | Yes | 36 | 70.6 |
|  | No | 11 | 21.6 |
|  | Unsure | 3 | 5.9 |
|  | No answer | 1 | 2.0 |
| Do you record and evaluate readmission rates? | Yes | 24 | 47.1 |
|  | No | 22 | 43.1 |
|  | Unsure | 3 | 5.9 |
|  | No answer | 2 | 3.9 |
| Which of the patient's sociodemographic characteristics below are recorded and used for evaluating potential inequalities in care? Please select all that apply | Ethnicity | 29 | 56.9 |
|  | Gender | 36 | 70.6 |
|  | Sex | 29 | 56.9 |
|  | Socioeconomic Status | 18 | 35.3 |
|  | Disability | 13 | 25.5 |
|  | Language | 13 | 25.5 |
|  | Religion | 14 | 27.5 |
|  | Other (please describe | 21 | 41.2 |
